# The effect of adult psychological therapies on employment and earnings: Evidence from England

**DOI:** 10.1101/2025.01.16.25320685

**Authors:** Klaudia Rzepnicka, Emma Sharland, Marta Rossa, Ted Dolby, Ekaterina Oparina, Rob Saunders, Daniel Ayoubkhani, Vahé Nafilyan

## Abstract

People suffering from common mental disorders (CMD) such as depression and anxiety are more likely to be economically inactive. Psychological therapies are highly effective at treating CMDs, but less is known about their impact on long-term labour market outcomes. Using national treatment programme data in England, NHS Talking Therapies (NHSTT), with unique linkage to administration data on employment and census records, we estimated the causal effects of NHSTT on employment and earnings.

Overall, completing treatment led to a maximum average increase of £17 in monthly earnings (year two) and likelihood of paid employment by 1.5 percentage points (year seven). Those “Not working, seeking work’ saw a maximum average increase in pay of £63 per month (year seven) and likelihood of paid employment by 3.1 percentage points (year four). Our findings demonstrate the economic benefits of treating CMDs, and how investing in mental health can impact labour market participation.

## Introduction

Common mental disorders (CMD) such as depression and anxiety are among the most commonly reported conditions among people who are economically inactive (that is, neither working nor looking for work) due to long-term sickness across the world^1–5^. Individuals with poor mental health are more likely to be unemployed, experience longer durations of unemployment and long-term sickness absences, and have lower incomes and be at higher risk of poverty^3–6^. Previous reports have estimated poor mental health to costs the economy around £110 billion a year^7^, accounting for approximately 5% of the UK GDP in 2019^8^. The primary contributors of these costs are economic inactivity, high staff turnover, increased sick leave and presenteeism in the workplace. Increasing trends of economic inactivity and rising rates of mental illness point to a growing impact of mental ill-health on the economy^9^.

In recent years, governments across the world have made efforts to mitigate the adverse effects of CMDs such as increasing funding for mental health care and facilitating greater access to evidence-based psychological therapies^10,11^. In England, the National Health Service (NHS) expanded the NHS Talking Therapies (NHSTT, formerly known as Improving Access to Psychological Therapies) as part of the NHS Long Term Plan^12^. This large-scale programme offers evidence-based psychological therapies for adults with CMDs such as anxiety disorders or depression^13^. In addition to improving mental health symptoms, the NHS long term plan seeks to improve access to mental health support for people in work and to support those seeking work and retaining employment^14^. Increasing productivity was an anticipated by-product of improving mental health symptoms and was part of the initial plan for NHSTT^15^. Several studies have provided evidence that psychological therapies are associated with improved employment outcomes in the short term, but there is limited evidence on long-term effects. Psychological treatments delivered by NHSTT are associated with improved employment outcomes shortly after discharge, as well as reduced healthcare utilisation^16–20^. However, there is limited evidence around whether psychological therapies lead to improved employment outcomes that are sustained over time. Randomized controlled trial evidence from Norway’s PRomPT service (which is based on the NHSTT programme) shows an increase in personal income and rates of working without receiving state benefits during the five years post-treatment when compared to treatment as usual in primary care (Smith O, Clark D, Hensing G, Layard R, Knapstad M. Cost-benefit of IAPT Norway and effects on work-related outcomes and health care utilization: Results from a randomized controlled trial using registry-based data. Manuscript submitted for publication). In Spain, results of the PSicAP trial which evaluated the impact of transdiagnostic cognitive behavioural therapy versus routine primary care showed beneficial effects on personal income in the year after treatment^21^. In contrast, evidence from Denmark that was not based on IAPT, found no effect on labour market outcomes like employment^22^. To our knowledge, there are no England-based studies that evaluate the long-term labour market effects of NHSTT using national population data to evaluate individual-level payment records. Having estimates of long-term effects of the NHSTT on employment and earnings would be crucial to provide inputs for cost-benefits calculations. Additionally, it will provide evidence supporting the increased spend in NHSTT committed by the UK government to get people who have a mental health condition back into work^23^. Whilst the NHSTT in England has been found to be cost-effective – with every pound spent, the programme generates an additional benefit of £5.50 for the economy – these calculations are based on extrapolating the immediate and short-term effects of therapy^24^.

In this study, we first aim to evaluate the effects of NHSTT treatment completion on labour market outcomes such as employment and earnings. Second, we aim to test whether the effect of treatment completion on labour market outcomes vary across different socio-demographic and treatment related characteristics. Third, using a parallel analysis we aim to evaluate the effects of positive treatment outcomes on the labour market.

## Methods

### Study design

Using a unique data linkage between the 2011 Census in England & Wales, His Majesty’s Revenue and Customs’ (HMRC’s) Pay As You Earn (PAYE) records, Office for National Statistics (ONS) death registrations and the NHSTT national dataset, we conducted a retrospective, quasi-experimental, panel-data study to evaluate the effects of psychological therapies on labour market outcomes (probability of being a paid employee and monthly employee earnings). Ethical approval was obtained from the National Statistician’s Data Ethics Advisory Committee (NSDEC23(18)).

### Data sources

The 2011 Census is a population-based survey conducted by the ONS to collect information to help build a snapshot of people and households in England and Wales (estimated response rate = 93.9%)^25^. Census data were linked to electronic health records from the NHSTT programme, a primary mental health service in England which offers evidence based psychological therapies for people with CMDs^26^. In 2021/22, 1.81 million referrals were received by NHSTT services, of which 1.24 million patients accessed the treatment^27^. All services across England collect standardised measures contributing towards the NHSTT Minimum Data Set (MDS). These data are processed into monthly and annual submissions by NHS England^28,29^. We obtained annual adult NHSTT data from NHS England via the Data Access Request Service (DARS).

Labour market outcomes were derived from the HMRC PAYE dataset which is collected for tax purposes in the UK. This dataset stores information submitted by employers on their employees’ salaries, bonuses, payrolled expenses, redundancy payments or allowances^30^. The dataset covers the whole employee population in the UK, excluding people who are self-employed or receive income from other sources such as investments or property rentals.

### Data linkage

NHSTT records were linked to the 2011 Census and death registrations via the NHS number. The 2011 census was linked to the 2011-2013 Patient Registers to retrieve NHS number for Census respondents, with a linkage rate of 95.3%^31^.

The HMRC PAYE data was linked via the encrypted National Insurance (NI) number. To obtain encrypted NI numbers, the census records were indexed to the ONS Demographic Index^32^ data via NHS number, with 89.7% of census records successfully linked to encrypted NI numbers. All datasets were de-identified before analysis.

### Data structure

Employees can be paid via the PAYE system according to a variety of payment frequencies: annual, quarterly, monthly, four-weekly, weekly, or irregularly. Payments were therefore calendarized such that the resulting linked dataset had a panel data structure, with monthly records for individuals. Where an individual had multiple monthly PAYE records (e.g. due to multiple employments), the pay recorded in the PAYE dataset was summed across all matching records for each month. Monthly records were aggregated into quarters to reduce the dataset size. Our final linked dataset had a panel structure with quarterly records for individuals. The quarters were defined using calendar months: January to March (Q1), April to June (Q2), July to September (Q3) and October to December (Q4).

### Study population

Our study population includes individuals with an NHSTT referral from 01 April 2016 to 31 March 2020, who attended at least one therapy session (that was not an assessment) and were scoring in the clinical range for either depression or an anxiety disorder^33^ at the time of referral. Participants were included in the sample when the reason for the termination (as reported by the therapist) was 1) suitable for NHSTT, but the patient refused the treatment that was offered 2) the patient completed scheduled treatment 3) the patient dropped out of treatment (unscheduled discontinuation) (**Supplementary Table 10**). Participants must also have been enumerated in 2011 Census and have valid NHS and NI numbers required for complete linkage. The population was restricted to those aged 25-60 years and living in England at the time of referral. This age restriction was implemented as younger adults (18-24 years) are likely to be in full-time education or moving into employment, therefore any effects seen in this analysis may likely be driven by natural movement into the workforce (see sensitivity analysis). Additionally, in selecting 25 years as a minimum age, we were able to have fuller pre-follow up history for all individuals within the analysis. We removed participants who died in the same calendar quarter as the referral and those who were retired when they were referred to NHSTT. For participants with multiple referrals, we selected the earliest one.

### Exposure variable

To evaluate effects of treatment completion, the exposed group comprised participants who were rated by their therapists as having completed treatment. The non-exposed group comprised participants who were suitable for NHSTT but refused the treatment that was offered, and participants who were rated by their therapist as having dropped out of treatment.

The main exposure was the time since the first therapy (excluding the initial assessment) for people who completed the treatment. Time period 0 refers to the date (year and quarter) in which the first therapy occurred; all modelled estimates of changes in pay and the likelihood of employment are relative to this reference period.

### Outcome variables

Two outcomes were analysed: monthly employee pay and paid employee status (binary). Monthly employee pay was derived by dividing the gross (pre-tax) quarterly pay recorded in PAYE by three. Monthly pay was imputed to be zero if it was negative, winsorized at the 99.9% centile to remove erroneously large values, and deflated to 2023 prices using the Consumer Price Index including owner occupiers’ housing costs (CPIH). Being a paid employee was defined as receiving any monthly pay greater than zero. For the pay outcome, analyses were conducted on both the full dataset, and a dataset including only the months for which individuals were a paid employee. Participants who had an NI number but no records in the HMRC PAYE dataset (for example, due to unemployment, long-term sickness or self-employment) were included in the analysis, and their earnings were set to 0.

### Covariates

Sociodemographic characteristics including age (calculated based on date of birth measured in quarter-year), country of birth (UK or non-UK), number of dependent children, relative area deprivation, disability, ethnicity, highest qualifications, National Statistics Socio-Economic Classification (NS-SEC), region, and sex were derived from 2011 Census (See **Supplementary Table 11**). Date of death was derived from death registrations and was used to censor the follow-up time of participants who died during the reporting period.

Diagnosis, employment status, psychotropic medication use, number of treatment sessions, source of referral, therapy intensity and treatment outcomes were taken from NHSTT data. We used the recorded information at the beginning of therapy for self-reported employment status and medication use. Treatment outcomes were predefined by NHSTT and are based on scores from Patient Health Questionnaire (PHQ-9) and Generalised Anxiety Disorder (GAD-7) (or an Anxiety Disorder Specific Measure (ADSM)) taken at the beginning and end of therapy. We derived four treatment outcomes (reliable recovery, reliable improvement, reliable deterioration, and no change) as per agreed NHS metrics^13^ and combined these into 1) reliable recovery or reliable improvement versus 2) reliable deterioration or no change.

### Statistical analysis

We assessed imbalance in the covariate distributions between the two treatment groups. Previous research indicates that absolute standardised mean differences <10% demonstrates balance between the treatment groups^34^. To help create a non-exposed group that was similar to the exposed group in terms of observed characteristics, we fitted a logistic regression including characteristics exhibiting the largest between-group imbalance to estimate the probability of completing treatment. We then used the predicted probability to calculate propensity scores and inverse probability weights (IPWs) to account for the differences in characteristics and thus mitigate against pre-treatment trends in outcomes. The variables with a >10% difference were: age band (25-34, 35-44, 45-54 and 55-60), number of dependent children, relative area deprivation, employment status at referral, highest qualification, NS-SEC and region (of residence). Disability was not included despite the standardised difference being 12.1% as the inclusion of this variable made little difference to monthly employee pay and employee status. We also did not include treatment outcome and number of treatment sessions as they are post treatment variables and therefore may be directly correlated with the outcome measures.

For the main analysis, we used an event study approach. The estimation equation (equation (1)) modelled the outcomes for individual *i* observed at time *t* (measured in quarter-years) and was fitted using linear regression with individual fixed effects (*α*_*i*_) to capture time invariant confounding factors. Fixed effects for each year since the first therapy (∑_*k*_ *γ*_*k*_ *Time*_*since*_*therapy*_*i*,*t*_), from four years prior to start to seven years after the start, were included to capture natural recovery. The exposure of interest was the interaction between time since the first therapy session and a time-invariant indicator variable for having completed the treatment (or positive treatment outcome), with time 0 being excluded. The models were adjusted for age and calendar time (quarter and year) to capture age-related employment progression and changes in background macroeconomic conditions. Participants’ follow-up was censored at the earliest of turning 64 years old, death, and end of study (31/12/2022).

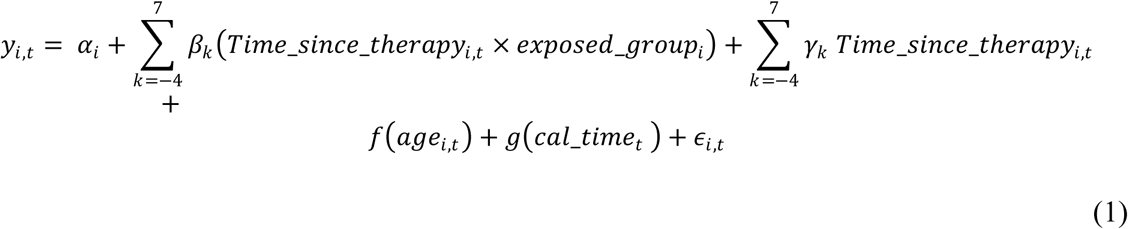

All reported confidence intervals around point estimates are at the 95% level. Statistical significance was determined by p < 0.05. Analysis of labour market outcomes by completing or dropping out of treatment was stratified by: age band, deprivation, diagnoses, employment status at referral, ethnicity, highest qualifications, psychotropic medication usage, NS-SEC, region, sex and therapy intensity. The parallel analysis investigating the effects of positive treatment outcomes was stratified by: age band and number of treatment sessions.

The initial processing of HMRC PAYE data was performed in Python version 3.6.8. Processing and analysis of the linked NHSTT data, Census 2011 data and death registrations were conducted in R version 4.1.3. All data were de-identified before being analysed in the secure data environment at the ONS.

### Heterogeneous treatment effects estimation

We first looked at heterogenous treatment effects by sociodemographic characteristics and labour market status, before applying a Generalized Random Forest (GRF) approach^35^ to study treatment heterogeneity effects without pre-selecting its sources. To adapt the model for GRF, we made two adjustments. First, since GRF estimates a single effect at a time, we focused on a single year. Year four was selected post-treatment to balance an ample follow-up period with a sufficient sample size. This subsample includes 630,991 observations, representing 75.0% of the total sample in the main analysis.

Next, as the GRF model does not incorporate covariates in the same way as traditional regression, we did not include any individual fixed effects. We transformed the outcome to reflect the change from the average level one year before treatment to the average four years after, to maintain a within-person estimate. The parameter of interest is presented in equation (2).

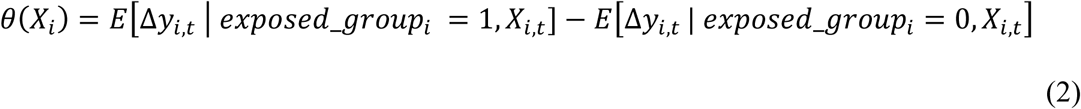

where Δ*y*_*i*,*t*_ = (*ȳ*_*i*,*y*=*year*4_ – *ȳ_i,y_*_=*year*0_) is a change in the outcome *y*_*i*_ of an individual *i* who had their first therapy session at time t, and *X*_*i*_ is a set of covariates.

The GRF creates multiple ‘decision trees,’ each of which sequentially splits the data into bins of observations that are similar within the bin but distinct from other bins in terms of the treatment effect. The treatment effect is then estimated for each bin. By sampling random subsets of data to build each tree and averaging the estimates for each individual across trees, GRF avoids overfitting and captures general patterns rather than any single tree’s result. In practice, the algorithm also uses different subsamples for growing trees and treatment effect estimation. This is known as the honest approach and helps reduce the risks of overfitting and biased estimates^24^. A histogram of individual estimates illustrates the range and variability of effects across the population. The algorithm also provides an overall average treatment effect.

In GRF, each decision tree split is based on a covariate, e.g. age being above or below a threshold. The *variable importance score* helps identify which covariates are most related to the differences in treatment effects by tracking how frequently each variable is used for splits, with greater weight given to splits earlier in the tree.

Like the parametric model above, the GRF accounts for the fact that this analysis, both the outcome and exposure likelihood can be partially explained by covariates. This was done by ‘orthogonalizing’ the forest: we first estimated individual propensity scores and outcomes using regression forests. We then calculated the residuals for the exposure and outcome variables and estimate a causal forest on these residuals. See Nie and Wager (2021) for details^36^.

The GRF algorithm was implemented with the `grf` package in R. The results are presented for 2,000 trees. To assess the sensitivity of the results, parts of the analysis were replicated for 500 trees. The results remained qualitatively similar.

### Sensitivity analysis

We conducted several sensitivity analyses. We re-ran the models swapping our exposed group to those participants who had either reliably recovered or improved. The non-exposed group comprised participants who had no change or deterioration of their mental health symptoms following NHSTT treatment. For this analysis participants with missing treatment outcomes were removed (N=5,000).

We also conducted analysis on monthly records for a random sample of participants (due to processing restrictions) to account for potential seasonal patterns masked by aggregated quarterly data. Additionally, to examine the extent to which age (at the time of referral) affected the labour market effects, we tested models with different inclusion criteria for age (18 years old at the time of referral with prior follow-up back to 16 years).

## Results

### Patient demographics

Of the 842,127 participants (mean age 40.5 years, s.d. 10.1) included in the analysis (see **Fig.1**), 593,300 (70.5%) were rated by their therapist to have completed treatment during the study period, and 248,827 (29.5%) were referred and assessed but refused the treatment that was offered or were rated by their therapist as having dropped out of treatment. The study population predominantly comprised: female (66.6% for the completed treatment group and 67.8% for the dropped out of treatment group), from a white ethnic background (90.2% and 89.7% respectively), who self-referred into NHSTT (74.1% and 72.6% respectively) and with primary diagnosis of depression (38.0% and 38.9% respectively) or generalized anxiety disorder (24.5% and 23.2% respectively). There were notable differences between groups across many characteristics, with the largest being for medication use (standardised mean difference 38.5%), qualifications (37.2%) and relative area deprivation (30.0%). See **Extended data Table 1** and **Extended data Table 2** for the analytical cohort descriptives.

**Figure 1.**
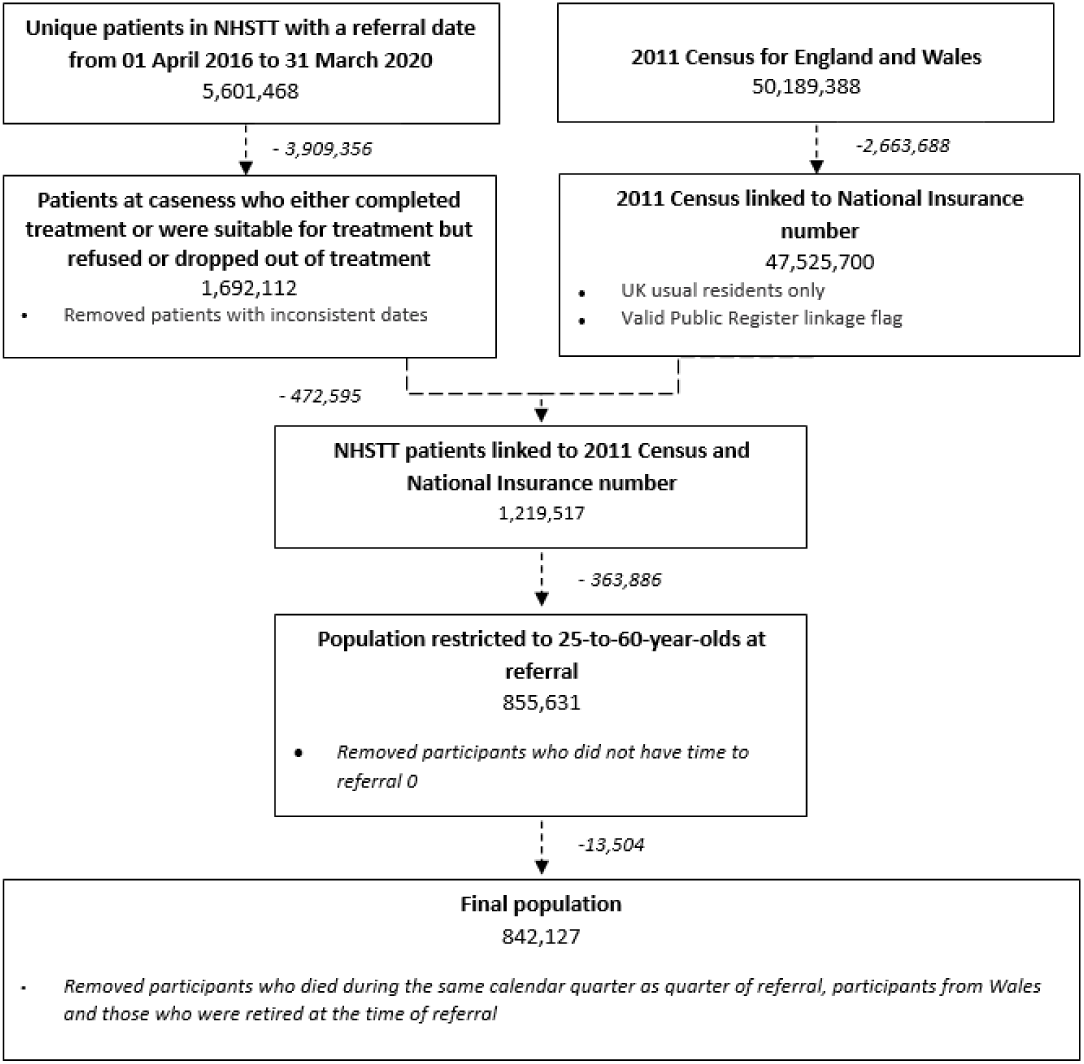
Population and linkage flow diagram.

For the exposed group, mean follow-up was 15.2 calendar quarters before starting therapy (out of maximum of 25 quarters) and 18.7 quarters after first therapy (maximum 26 quarters). For the non-exposed group, mean follow-up was 15.3 quarters before starting therapy and 18.6 quarters after therapy **(Supplementary Table 1).** On the date of the first therapy (quarter and year), 68.7% of individuals were in paid employment (70.9% for exposed and 63.4% for non-exposed), while the mean monthly deflated earnings was £1,615.0 (£1,716.6 exposed and £1,372.9 non-exposed) when including those not in work, and £2,351.79 (£2,421.6 exposed and £2,165.6 non-exposed) when excluding those not in work (**Supplementary Table 2**). The exposed group earned more on average than the non-exposed group before and after treatment, while the IPWs made the earnings trajectories more parallel (**Supplementary Figure 1**).

### Effect of treatment completion on monthly earning and employee status

The effect of NHSTT on monthly earnings reached a maximum of £16.9 more per month [95% CI 11.6-22.2] in year two after treatment, and the statistically significant increase relative to pre-treatment was sustained at £10.6 [95% CI 0.2-21.0] six years after treatment. Though the magnitude of the effect remained similar in year seven to year six, the confidence intervals indicated the results were no longer statistics significant due to diminished power (**Fig.2**). We also found statistically significant evidence that completing NHSTT is associated with an increase in the probability of being a paid employee; the maximum effect reached and was sustained at 1.5 p.p. [95% CI 1.0-2.0] seven years after first therapy (**Fig.2**).

**Figure 2.**
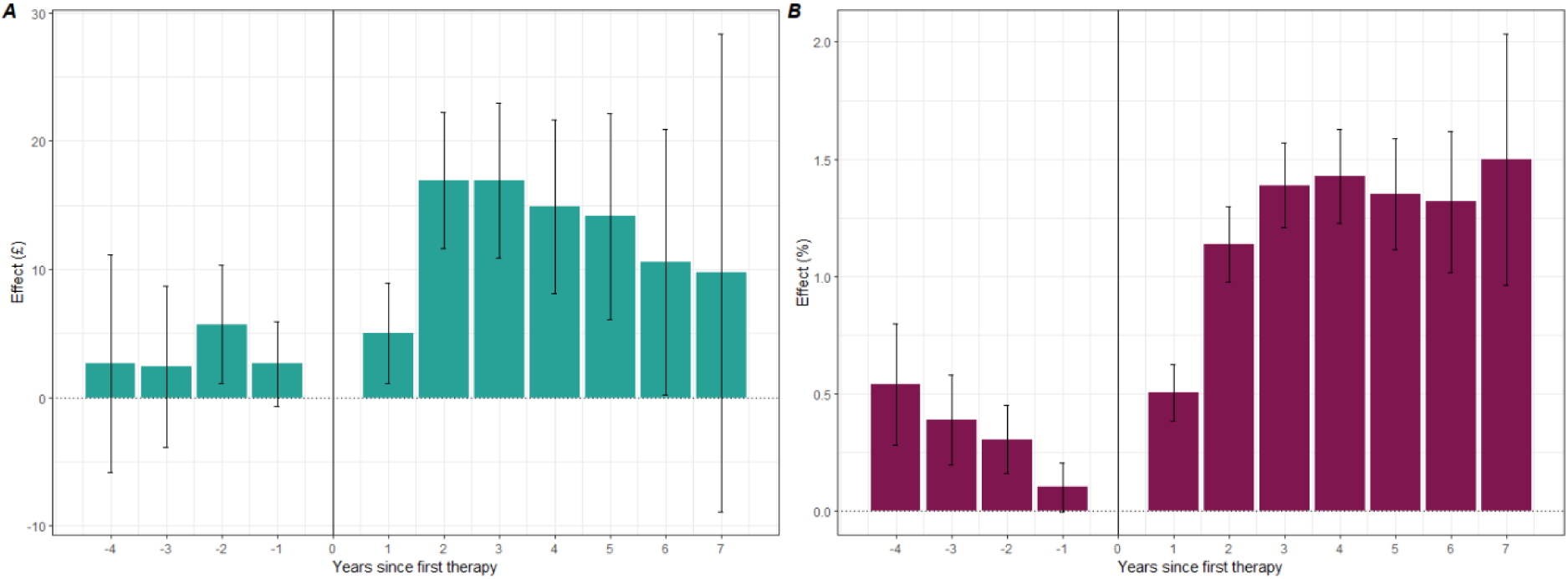
Effects of NHSTT talking therapies treatment completing on (A) monthly earnings and (B) probability of being a paid employee.

### Heterogenous treatment effects by sociodemographic characteristics

Participants who reported to be ‘Not working, seeking work’ at beginning of therapy saw greatest increases in the probability of being employed and average monthly earnings; compared with the one year before first therapy, the effect on monthly earnings reached maximum of £63.0 [95% CI 17.7-108.3] more per month by year seven after treatment. Probability of employment reached maximum of 3.1 p.p. [95% CI 2.4-3.9] in year four and the increase was sustained at 3.0 p.p. [95% CI 1.3-4.7] by year seven (**Fig.3**). The effect size of the NHSTT treatment completion was smaller but still positive for the ‘Not working, not seeking work’; 1.5 p.p. [95% CI 0.7-2.3] four years after therapy (largest year effect) and 0.5 p.p. [95% CI −1.5-2.5] in year seven (there was no statistically significant effect on monthly earnings in this group).

**Figure 3.**
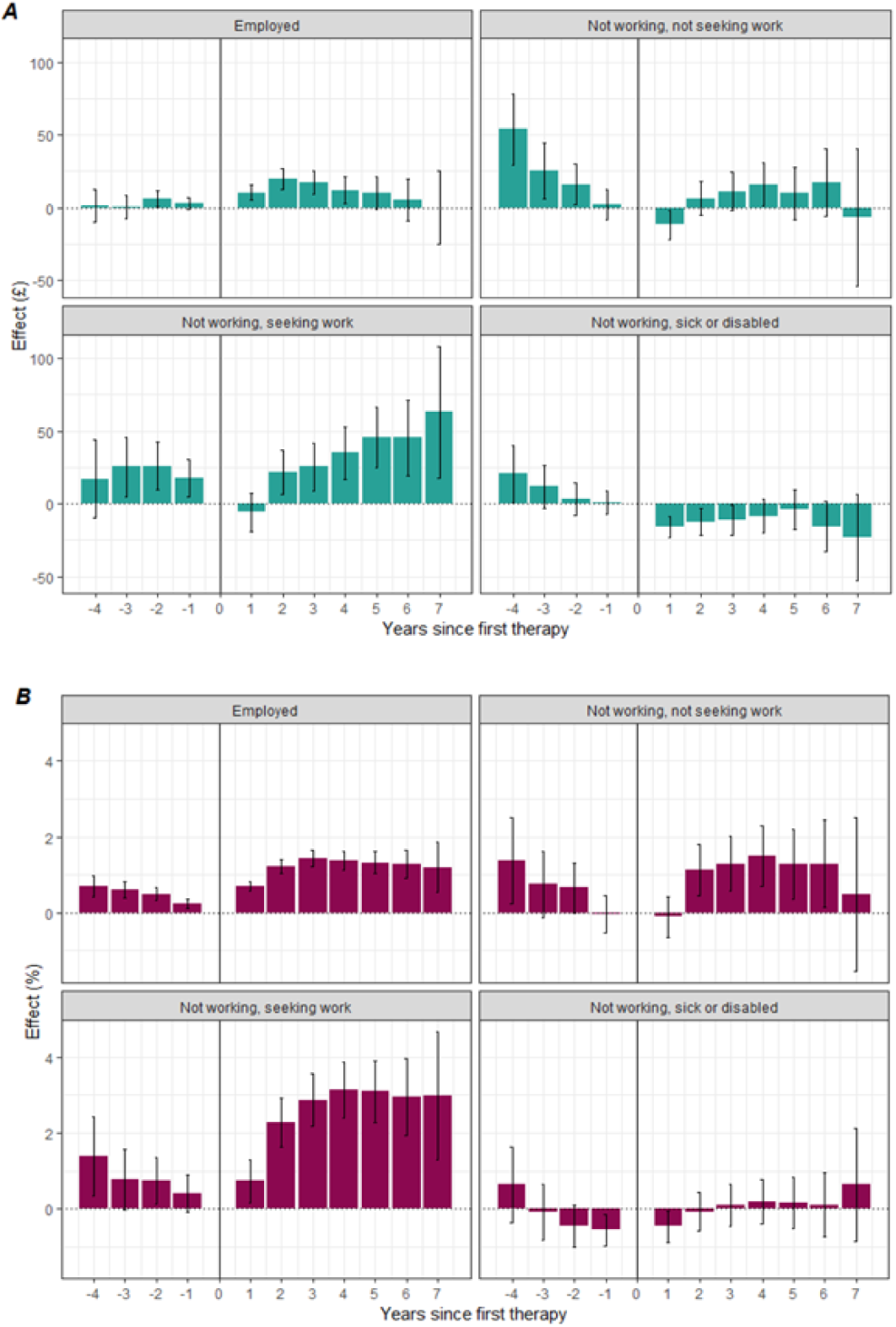
Effects of NHSTT treatment completion on (A) monthly earnings and (B) probability of being a paid employee broken down by self-reported employment status (recorded at the beginning of therapy).

For both females and males, monthly earnings were highest in year three after treatment (£15.9 [95% CI 9.3-22.5] for females and £24.8 [95% CI 12.4-37.2] for males). The probability of being a paid employee was highest in year seven after treatment; compared with the year of first therapy, the effect reached 1.4 p.p. [95% CI 0.5-2.3] for males and 1.5 p.p. [95% CI 0.9-2.2] for females (**Supplementary Table 3 and 4**).

Overall, completing NHSTT treatment had the largest effect in the age groups 25-34 and 35-44 years; the effect reached maximum of 2.3 p.p. [95% CI 1.4-3.2] for the 25-34-year-olds (in year seven) and 2.0 p.p. [95% CI 1.6-2.4] for the 35-44-year-olds (in year five) (**Supplementary Table 4**). Results for 45-54-year-olds were smaller but still positive, with the highest probability reaching 1.0 p.p. [95% CI 0.6-1.3] in year three. By year seven after treatment, the increase relative to pre-treatment was sustained at 1.9 p.p. [95% CI 0.9-2.8] for 35-44-year-olds, and 1.0 p.p. [95% CI 0.0-2.0] for 45-54-year-olds. Completing NHSTT treatment had no statistically significant effect on individuals aged 55-60 years. See **Supplementary Table 3** for effect on monthly earnings. See **Supplementary Table 3** and **Supplementary Table 4** for further sociodemographic and treatment characteristic breakdowns.

Using the GRF approach, for both outcomes, monthly earnings and probability of being a paid employee, we found heterogeneity for respondents with different characteristics four years after therapy. Neither of the two estimated distributions was bounded away from zero, implying that there are groups of patients who do not benefit from treatment (**Supplementary Figure 2**). This is supported by results from **Fig.3A**. which show that the main benefits of NHSTT are in population ‘Not working, seeking work’. Additionally, when restricting the population across the study period to individuals in paid employment only (pay>0), the results from the fitted linear regression model with individual fixed effects found negative effects on monthly earnings of maximum of -£27.8 in year seven relative to one year pre-treatment (**Supplementary Table 5**). The conditional average treatment effect on the treated group was £23.2 (s.e.=12.0) for pay and 1.1 p.p. for probability of being a paid employee (s.e.=0.1). The top three variables identified as important for heterogeneity in probability of being a paid employee were age band (18.2%), ‘Not working, seeking work’ (14.3%) and ‘Students’ (10.2%). For monthly earnings, the top three variables were age bands (6.7%), quarter of first therapy (3.4%) and dependent children (3.1%). The importance of other variables can be found in **Supplementary Table 6**.

### Sensitivity analyses

The first part of the sensitivity analyses assessed the impact of swapping the exposure groups. The exposed group for this analysis includes individuals who showed a positive treatment outcome (N=645,381) and the non-exposed group includes individuals who had no change or reliable deterioration in their mental health symptoms (N=191,746) (See **Supplementary Table 7 and 8** for summary statistics). Positive treatment outcomes were strongly associated with improved monthly earnings and employee status. The largest effect on monthly earnings was found in year three after treatment and was equal to £49.7 [95% CI 43.2-56.2] more per month; the statistically significant increase relative to pre-treatment was sustained at £31.7 [95% CI 11.9-51.4] in year seven (**Fig.4**). Similarly, the probability of being a paid employee increased by 2.8 p.p. [95% CI 2.6-3.0] four years after therapy (largest effect). From year two onwards, the probability of being a paid employee was sustained and by year seven the increase reached 2.3 p.p. [95% CI 1.8-2.9] (**Fig.4**). For breakdowns by age and number of treatment sessions, see **Supplementary Table 9**.

**Figure 4.**
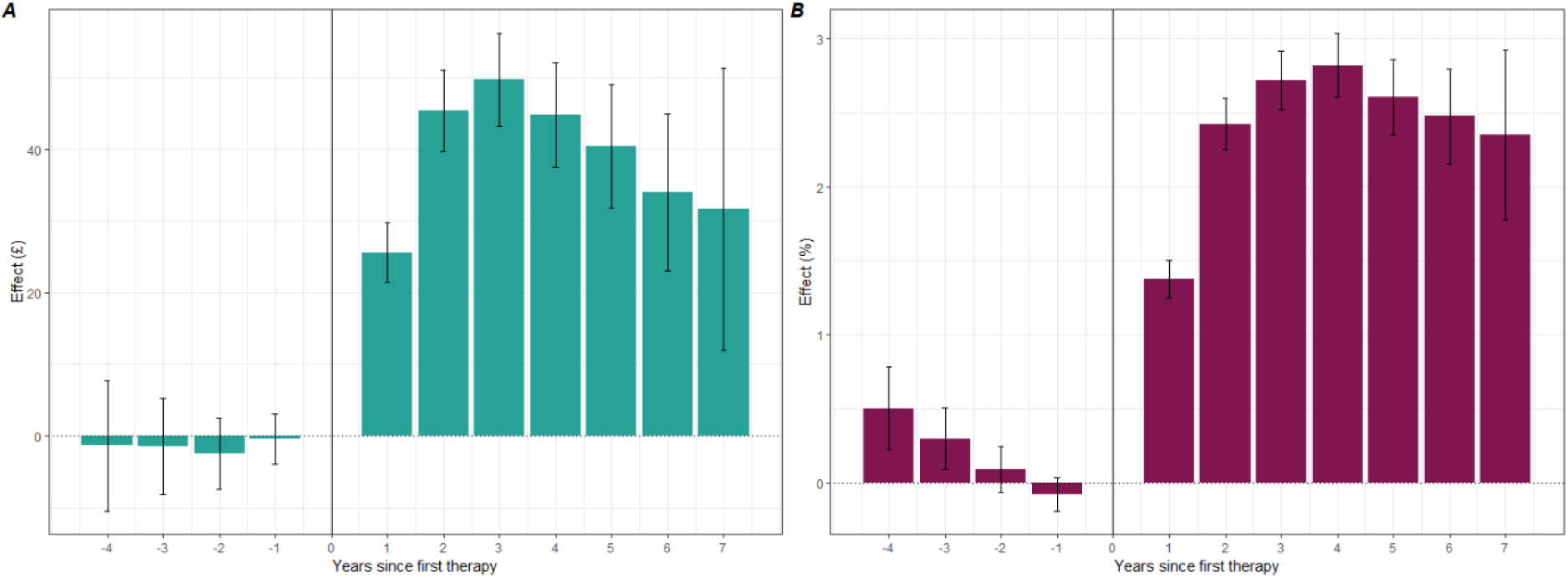
Effects of positive treatment outcomes after psychological therapies treatment on (A) monthly earnings and (B) probability of being a paid employee. The non-exposed group includes participants with no change or deterioration in their mental health symptoms.

The results from analysis on monthly records for a random sample of participants were in line with those from aggregated quarterly data (**Supplementary Table 10**). The effect size of the overall models was larger when we changed the inclusion criteria for age (18 years at time of referral with prior follow-up back to 16 years), likely driven by younger participants entering the workforce for the first time (**Supplementary Table 10**).

## Discussion

This paper is the first nationally representative analysis of long-term labour market effects of psychological therapies on employment and earnings globally. Our key findings show completing the NHSTT programme is associated with a sustained improvement in labour market outcomes: a 1.5 p.p. increase in the probability to be a paid employee seven years after treatment and a £10.6 average increase in monthly earning six years after treatment. A 1.5p.p increase in employment following treatment equates to around 10,000 people annually^(1)^.

Research has identified that mental ill-health is associated with an increased risk of economic inactivity and reduced wages, with some studies suggesting a potential causal link^37–39^. Those studies highlight the importance of psychological treatment for getting people into, or retaining them in, employment and increasing their earnings. Previous research has identified benefits of NHSTT treatment on labour market outcomes in the short term, with research suggesting between 4 and 10 percentage point increases in the employment rate over a maximum follow-up of 72 weeks^18,20^. Our research supports these initial studies confirming positive labour market outcomes of treatment in NHSTT. Whilst we see smaller overall improvements in the likelihood of being employed in our study, we demonstrate that these gains are sustained over the long-term post-treatment. We can also be confident that these increases are because of completing treatment due to our within-individual estimation, and the use of a non-exposed group of people who interrupted treatment earlier than expected.

Other studies have assessed benefits in the short-term using self-report methods, which are potentially subject to respondent bias, without suitable non-exposed groups for comparison^18,20^. Our findings are in line with research from low- and middle-income countries on effects of mental health interventions on labour market^40^.

The results from the data-driven GRF approach also support findings from the main analysis. Age band was highlighted as the most significant variable for both outcomes suggesting this variable is crucial in explaining the variations in employment outcomes. This is likely driven by the relationship between age and employment status during different stages of life^41^ as well as how well different groups respond to treatment. This might also reflect young people entering the workforce for the first time, salary and career progression driven by work experience, increased workplace stress over time^4^ or transition to retirement. Official statistics show that larger number of people aged 16 to 34 years are economically inactive due to depression, bad nerves, and anxiety (19% as compared to 14% for 35-49-year-olds and 9% 50-64-year-olds)^42^. We find probability of being a paid employee following NHSTT treatment to be highest in younger people, highlighting that targeting treatment for the groups mostly affected by CMDs^43^ can improve economic health of the country long-term.

Additionally, in this study we found no significant effects for individuals aged 55-60 years. A previous evaluation of psychological therapies found older people tend to respond better to treatment and reach higher recovery rates than other age groups^44^, they do not appear to have improved long-term employment outcomes. In line with our findings, a systematic review on mental health and retirement found that poor mental health is highly associated with early retirement in both men and women^45^. Therefore, improving mental health symptoms in older people could mitigate this risk and consequently improve employment outcomes for the individual in the long-term, as well as protecting their mental health by having stable and continuous employment^4^.

We found a decreasing trend in monthly earnings for those who self-reported as employed at the time of referral. Individuals who were unemployed (i.e. not working but seeking work) at the time of referral benefited the most from completing NHSTT treatment in terms of labour market outcomes. This characteristic was also identified as a meaningful contributor to heterogeneity in probability of being a paid employee. This may suggest that the effects of NHSTT on labour market outcomes are driven by the unemployed population moving into employment. The decreasing and negative effects on monthly earnings in the employed population might be explained by individuals reducing their working hours to facilitate improvements in mental health, avoid work related burn-out following a successful psychological therapy or moving to a job that makes them more happy but pays less^46,47^. Those finding support the idea that NHSTT can facilitate the unemployed to take up new jobs.

### Strengths and limitations

We recognise that our definition of completed treatment was based on a clinician-defined reason for the end of the care episode, which may be inconsistently applied. There is potential that the number of sessions attended, or whether positive benefit of treatments was reported, might impact recording of this variable. In addition, some of the patients who discontinued earlier than their therapist expected might have done so because they recovered with a smaller dose of therapy and did not come back to the service. The presence of such individuals in the non-exposed group could lead to underestimation of the magnitude of the effect of therapy on labour market participation.

The HMRC PAYE dataset is limited to people in paid employment; it excludes individuals who are self-employed or receive income from other sources, therefore, we cannot estimate the full extent of NHSTT on the labour market. However, official statistics report that in December 2022, the number of payrolled employees reached 29.9 million^30^ which suggests that our results are highly relevant for the majority of the working population. A potentially larger threat to the identification strategy is that the decision to discontinue treatment is endogenous, meaning that the patients who dropped out might be systematically different from those who remained in the treatment. The IPW addresses this in the best available way.

Lower income or probability of employment could be a positive outcome for an individual, especially if work is the direct cause of mental health issues (for example, due to being in an adverse psychosocial environment or working long hours). Future iterations of this research should look at other labour market outcomes such as sustained employment or receipt of social security benefits.

Lastly, our study is a non-randomized comparison. It has the advantage that it uses complete population data, but we may have under-estimated labour market effects. Consistent with this, the beneficial effects on income observed in our analyses are less than half those observed in the randomized controlled trial of the PRomPT service (Smith O, Clark D, Hensing G, Layard R, Knapstad M. Cost-benefit of IAPT Norway and effects on work-related outcomes and health care utilization: Results from a randomized controlled trial using registry-based data. Manuscript submitted for publication.). Taken together, our large scale non-randomized comparison and the much smaller randomized comparisons make a compelling case of the beneficial effects of NHSTT-style services on labour market participation.

### Conclusions

Completion of psychological treatment for CMDs through the national NHSTT programme leads to sustained increases in both employment and earnings up to seven years after the start of treatment, equating to approximately 10,000 people back into employment annually. Policymakers may consider ways to invest in mental health services such as NHSTT to reduce the impact of CMDs on the economy. Future research should investigate the impact on labour market outcomes of physical comorbidities in addition to mental health conditions, explore a broader range of outcomes such as social security benefit receipts, and consider the implications of the employment advisors service provided by NHSTT to understand in more detail the mechanisms underpinning our findings.

## Data availability

The source data are not publicly available and are subject to controlled access due to their sensitive nature. Census 2011 and death registration data are available through the Integrated Data Service (IDS). Details of the application requirements and process, and the use of data, are available at https://integrateddataservice.gov.uk/how-to-access-the-integrated-data-service. The NHS Talking Therapies dataset is held by NHS England and can be accessed through the NHS Secure Data Environment: https://digital.nhs.uk/services/secure-data-environment-service.

## Code availability

The code for the project is available here: https://github.com/ONS-Health-modelling-hub/eafnhstt_project_phase1.git

## Acknowledgements

This project was funded by the Cabinet Office and HM Treasury Evaluation Task Force’s Evaluation Accelerator Fund (2023 to 2024) and by HM Treasury’s Labour Markets Evaluation and Pilots Fund (2024 to 2025).

## Author contributions

ES and VN conceived, acquired funding and designed the study. KR led the data analysis with support from; VN, MR, EO and TD. MR and KR undertook quality assurance of the code and data. ES, VN, DA, KR, RS and EO quality assured the outputs and the interpretation of them. ES, KR, VN, DA, RS and EO contributed to critical revision of the manuscript and approved the final manuscript.

## Supplementary Information

Supplementary Information is available for this paper. Supplementary Figure 1-2 and Tables 1-11.

## Extended data tables

**Extended data table 1.**
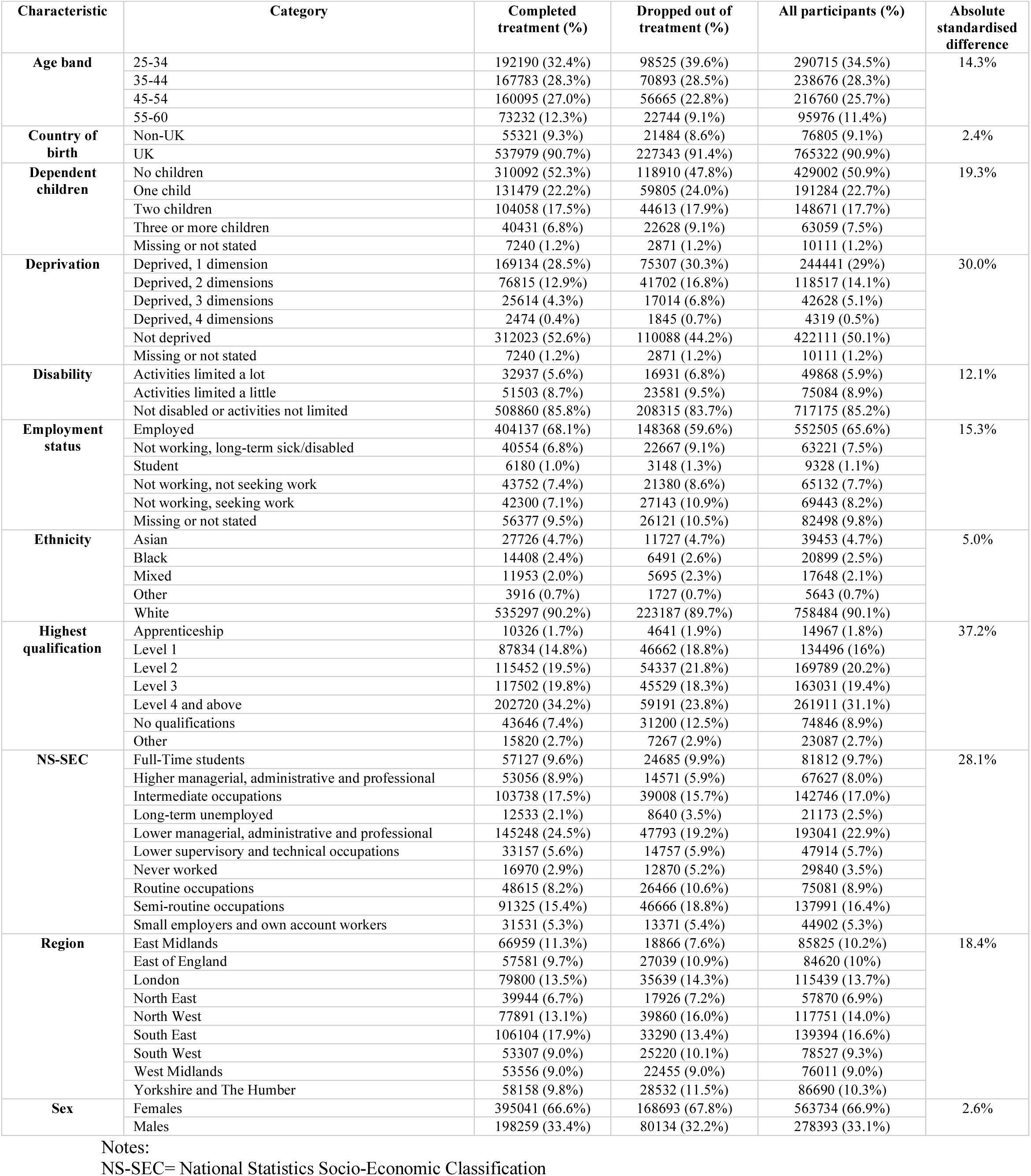
Sociodemographic summary statistics between the completed and dropped out groups.

**Extended data table 2.** Mental health and treatment summary statistics between the two treatment groups.

| Characteristic | Category | Completed treatment (%) | Dropped out of treatment (%) | All participants (%) | Absolute standardised difference |
| --- | --- | --- | --- | --- | --- |
| **Diagnosis** | Agoraphobia | 3166 (0.5%) | 2161 (0.9%) | 5327 (0.6%) | 8.0% |
|  | Depression | 225339 (38.0%) | 96887 (38.9%) | 322226 (38.3%) |  |
|  | Generalized anxiety disorder | 145633 (24.5%) | 57659 (23.2%) | 203292 (24.1%) |  |
|  | Mixed anxiety and depressive disorder | 60874 (10.3%) | 24298 (9.8%) | 85172 (10.1%) |  |
|  | Obsessive-compulsive disorder | 12774 (2.2%) | 4526 (1.8%) | 17300 (2.1%) |  |
|  | Other anxiety or stress related disorder | 26092 (4.4%) | 9261 (3.7%) | 35353 (4.2%) |  |
|  | Panic disorder | 14611 (2.5%) | 6983 (2.8%) | 21594 (2.6%) |  |
|  | Post-traumatic stress disorder | 22062 (3.7%) | 10033 (4.0%) | 32095 (3.8%) |  |
|  | Social phobias | 11805 (2.0%) | 5451 (2.2%) | 17256 (2.0%) |  |
|  | Specific (isolated) phobias | 4355 (0.7%) | 1556 (0.6%) | 5911 (0.7%) |  |
|  | Missing or not stated | 66589 (11.2%) | 30012 (12.1%) | 96601 (11.5%) |  |
| **Medication (Psychotropic Medication)** | Not prescribed | 253962 (42.8%) | 91263 (36.7%) | 345225 (41%) | 38.5% |
|  | Prescribed, not taking | 25601 (4.3%) | 12754 (5.1%) | 38355 (4.6%) |  |
|  | Prescribed, taking | 284327 (47.9%) | 132358 (53.2%) | 416685 (49.5%) |  |
|  | Missing or not stated | 29410 (5.0%) | 12452 (5.0%) | 41862 (5.0%) |  |
| **Number of treatment sessions** | 2 | 28387 (4.8%) | 72414 (29.1%) | 100801 (12.0%) | 111.3% |
|  | 3 to 5 | 154457 (26.0%) | 115087 (46.3%) | 269544 (32.0%) |  |
|  | 6 to 8 | 206771 (34.9%) | 38390 (15.4%) | 245161 (29.1%) |  |
|  | 9 to 15 | 149332 (25.2%) | 19228 (7.7%) | 168560 (20.0%) |  |
|  | 16 or more | 54353 (9.2%) | 3708 (1.5%) | 58061 (6.9%) |  |
| **Source of referral** | Other | 34729 (5.9%) | 16466 (6.6%) | 51195 (6.1%) | 14.3% |
|  | Primary Health Care | 116870 (19.7%) | 51233 (20.6%) | 168103 (20.0%) |  |
|  | Self-referral | 439684 (74.1%) | 180624 (72.6%) | 620308 (73.7%) |  |
|  | Missing or not stated | 2017 (0.3%) | 504 (0.2%) | 2521 (0.3%) |  |
| **Therapy intensity** | High intensity | 397578 (67.0%) | 141379 (56.8%) | 538957 (64.0%) | 21.1% |
|  | Low intensity | 193951 (32.7%) | 106591 (42.8%) | 300542 (35.7%) |  |
|  | Missing or not stated | 1771 (0.3%) | 857 (0.3%) | 2628 (0.3%) |  |
| **Treatment outcomes** | Deterioration | 14929 (2.5%) | 22918 (9.2%) | 37847 (4.5%) | 201.1% |
|  | Improvement | 94193 (15.9%) | 79518 (32.0%) | 173711 (20.6%) |  |
|  | No change | 58830 (9.9%) | 95069 (38.2%) | 153899 (18.3%) |  |
|  | Recovery | 424142 (71.5%) | 47528 (19.1%) | 471670 (56.0%) |  |
|  | Missing or not stated | 1206 (0.2%) | 3794 (1.5%) | 5000 (0.6%) |  |

## Footnotes

(1) In 2023/24, 1.8 million people were referred to NHSTT, of whom 671,648 completed treatment^48^.

